# Robust methylation-based classification of brain tumors using nanopore sequencing

**DOI:** 10.1101/2021.03.06.21252627

**Authors:** Luis P. Kuschel, Jürgen Hench, Stephan Frank, Ivana Bratic Hench, Elodie Girard, Maud Blanluet, Julien Masliah-Planchon, Martin Misch, Julia Onken, Marcus Czabanka, Philipp Karau, Naveed Ishaque, Elisabeth G. Hain, Frank Heppner, Ahmed Idbaih, Nikolaus Behr, Christoph Harms, David Capper, Philipp Euskirchen

## Abstract

DNA methylation-based classification of cancer provides a comprehensive molecular approach to diagnose tumors. In fact, DNA methylation profiling of human brain tumors already profoundly impacts clinical neuro-oncology. However, current implementations using hybridization microarrays are time-consuming and costly. We recently reported on shallow nanopore whole-genome sequencing for rapid and cost-effective generation of genome-wide 5-methylcytosine profiles as input to supervised classification using random forests complemented by a medium-resolution copy number profile derived from the same raw data. Here, we demonstrate that this approach allows to discriminate a wide spectrum of primary brain tumors using public reference data of 82 distinct tumor entities. We developed a pseudo-probability score as a confidence score for interpretation in a clinical context. Using bootstrap sampling in a discovery cohort of N = 56 cases, we find that a minimum set of 1,000 random CpG features is sufficient for high-confidence classification by ad hoc random forests for most cases and demonstrate robustness across laboratories with matching results in 13/13 cases. When applying the confidence score threshold to an independent validation series (N = 111), the method demonstrated 100% specificity for the remaining 93 cases. In a prospective benchmarking (N = 15), median time to results was 21.1 hours. In conclusion, nanopore sequencing allows robust and rapid methylation-based classification across the full spectrum of brain tumors. The integrated confidence score facilitates possible clinical implementation, while requiring further prospective evaluation.

## Introduction

DNA methylation is a stable epigenomic mark of cell identity and has become a powerful tool in tissue-based cancer diagnosis. Application of supervised machine learning to genome-wide DNA methylation profiles enables classification of unknown sample with respect to a reference cohort, providing a method that is both independent and complementary to histomorphology. It has been used for a variety of pan-cancer classification approaches, such as brain tumors [1], soft tissue sarcoma [7] or cancer of unknown primary (CUP) [12]. In addition to offering an unbiased method of mandatory molecular testing, required by the current WHO classification of CNS tumors (e.g., for medulloblastoma subtypes), systematic application of methylation-based classification has revealed weaknesses of histomorphology, which is associated with misdiagnoses in approximately 10% of cases [1]. However, for current microarray-based implementations, turnaround times are in the range of several days to weeks [6] due to complex wet-lab procedures and capital cost, multiplex assays allow reasonable per-assay cost only in high-throughput settings like tertiary care centers.

To address these shortcomings, we have recently used nanopore whole-genome sequencing for simultaneous generation of copy number and DNA methylation profiles [2]. Nanopore sequencing determines DNA sequence as well as base modifications, such as 5-methylation of cytosine (5mC), by detecting changes in ionic currents when molecules pass a biological nanopore inserted in a dielectric membrane [13,16]. Methylation detection in native DNA in combination with time-efficient hands-on procedures in the range of less than an hour in total significantly decreases turnaround times.

Here, we extend this pilot approach to methylation-based classification of 82 brain tumor entities, define criteria for robust diagnostic implementation and provide benchmarking data from cross-laboratory as well as cross-method validation. Moreover, we share first insights into prospective analysis of turnaround times in comparison to routine diagnostic procedures as part of a planned multicentric clinical trial.

## Methods

### Experimental design

In order to refine machine learning and to define quality criteria for reliable classification, a discovery cohort (N = 56) was compiled, containing 15 prospective cases and 41 retrospective cases from previously published datasets [2,15]. The local ethics committee (Charité – Universitätsmedizin Berlin, Berlin, Germany; EA2/041/18) approved generation of prospective data in the context of this study. Robustness of classification was then assessed in an independent validation cohort (N = 111) generated during routine clinical application at the Division of Neuropathology, Institute of Pathology, Basel, Switzerland (as an in-house validated diagnostic test) (Suppl. Fig.1). All cases underwent routine diagnostic procedures, including microarray-based analysis [1] in 109/111 cases, and were classified in accordance with the 2016 World Health Organization Classification of Tumors of the Central Nervous System [10] (Table 1).

**Table 1.** Summary of tumor entities in this study. Reference diagnosis is reported in accordance with the 2016 WHO classification of central nervous system tumors. A detailed summary of clinical characteristics of all cases can be found in Supplementary Table 1.

|  | discovery<br>cohort<br>(N=56) | validation<br>cohort<br>(N=111) | Overall<br>(N=167) |
| --- | --- | --- | --- |
| **WHO integrated diagnosis** |  |  |  |
| Anaplastic astrocytoma, IDH-mutant | 2 (3.6%) | 3 (2.7%) | 5 (3.0%) |
| Anaplastic oligodendroglioma, IDH-mutant and 1p/19q-codeleted | 3 (5.4%) | 1 (0.9%) | 4 (2.4%) |
| Angiomatous meningioma | 1 (1.8%) | 0 (0%) | 1 (0.6%) |
| Atypical teratoid/rhabdoid tumour | 3 (5.4%) | 0 (0%) | 3 (1.8%) |
| CNS embryonal tumour, NOS | 1 (1.8%) | 0 (0%) | 1 (0.6%) |
| CNS Ewing sarcoma family tumor with CIC alteration | 1 (1.8%) | 0 (0%) | 1 (0.6%) |
| CNS neuroblastoma with FOXR2 activation | 1 (1.8%) | 0 (0%) | 1 (0.6%) |
| Diffuse astrocytoma, IDH-mutant | 2 (3.6%) | 1 (0.9%) | 3 (1.8%) |
| Diffuse midline glioma, H3 K27M-mutant | 1 (1.8%) | 0 (0%) | 1 (0.6%) |
| Embryonal tumour with multilayered rosettes, C19MC-altered | 1 (1.8%) | 1 (0.9%) | 2 (1.2%) |
| Glioblastoma, IDH wildtype, H3.3 G34 mutant* | 1 (1.8%) | 0 (0%) | 1 (0.6%) |
| Glioblastoma, IDH-mutant | 2 (3.6%) | 1 (0.9%) | 3 (1.8%) |
| Glioblastoma, IDH-wildtype | 26 (46.4%) | 24 (21.6%) | 50 (29.9%) |
| Medulloblastoma, genetically defined, non-WNT/non-SHH | 4 (7.1%) | 0 (0%) | 4 (2.4%) |
| Medulloblastoma, genetically defined, WNT-activated | 2 (3.6%) | 0 (0%) | 2 (1.2%) |
| Medulloblastoma, NOS | 1 (1.8%) | 0 (0%) | 1 (0.6%) |
| Medulloblastoma, SHH-activated | 1 (1.8%) | 0 (0%) | 1 (0.6%) |
| Meningothelial meningioma | 2 (3.6%) | 0 (0%) | 2 (1.2%) |
| Pleomorphic xanthoastrocytoma | 1 (1.8%) | 0 (0%) | 1 (0.6%) |
| Diffuse large B cell lymphoma (DLBCL) of the CNS | 0 (0%) | 3 (2.7%) | 3 (1.8%) |
| Ependymoma | 0 (0%) | 3 (2.7%) | 3 (1.8%) |
| Fibrous meningioma | 0 (0%) | 1 (0.9%) | 1 (0.6%) |
| Glioblastoma, IDH wildtype, subclass midline* | 0 (0%) | 1 (0.9%) | 1 (0.6%) |
| Haemangioblastoma | 0 (0%) | 1 (0.9%) | 1 (0.6%) |
| Medulloblastoma, genetically defined, group 4 | 0 (0%) | 1 (0.9%) | 1 (0.6%) |
| Medulloblastoma, genetically defined, SHH-activated and TP53-wildtype | 0 (0%) | 1 (0.9%) | 1 (0.6%) |
| Meningioma | 0 (0%) | 49 (44.1%) | 49 (29.3%) |
| Pilocytic astrocytoma | 0 (0%) | 3 (2.7%) | 3 (1.8%) |
| Pituitary adenoma ACTH producing | 0 (0%) | 2 (1.8%) | 2 (1.2%) |
| Pituitary adenoma gonadotropin producing | 0 (0%) | 5 (4.5%) | 5 (3.0%) |
| Schwannoma | 0 (0%) | 8 (7.2%) | 8 (4.8%) |
| Solitary fibrous tumour / haemangiopericytoma | 0 (0%) | 1 (0.9%) | 1 (0.6%) |
| Subependymal giant cell astrocytoma | 0 (0%) | 1 (0.9%) | 1 (0.6%) |

\* Of note, these entities are not yet recognized as distinct entities in the 2016 WHO classification.

### Patient material and tissue processing

Fresh tumor tissue of prospective cases was transferred to the local neuropathology laboratory during routine diagnostic procedures. Tissue was then snap-frozen and H&E cryosections were inspected to assess tumor purity. DNA was extracted using spin columns (DNeasy Blood & Tissue Kit, Qiagen, NL) according to the manufacturer’ s protocol with ∼25mg of tumor tissue. For the cross-laboratory cohort, tumor DNA obtained from the Institute Curie was extracted using phenol chloroform procedure. Eluted genomic DNA was quantified on a Qubit 4.0 fluorometer using the dsDNA BR Assay (Thermo Fisher, USA) and quality controlled using the 260/280 ratio (NanoDrop, Thermo Fisher, USA).

### Nanopore whole-genome sequencing

Library preparation with barcode labelling was performed with ∼400ng input of genomic DNA using the Rapid Barcoding Kit (RBK004, Oxford Nanopore Technologies, UK) according to manufacturer’ s instructions. During library preparation, input DNA is fragmented while simultaneously attaching barcodes using a time-efficient transposase-based approach. The final library was loaded onto a R9.4.1 flow cell (FLO-MIN106D, Oxford Nanopore Technologies, UK; alternatively, FLG-0001 cells sharing the architecture with FLO-MIN106D, were used) and whole-genome sequencing was performed for 6 to 24 hours on a MinION Mk 1B device (Oxford Nanopore Technologies, UK). FAST5 files containing the raw data were obtained in real-time using the manufacturer’ s software MinKNOW (v.1.3.1-v.3.6.0) and transferred to a high-performance computing (HPC) cluster for further analysis. Each flow cell was washed after sequencing (WSH002/WSH003, Oxford Nanopore Technologies, UK) and reused for up to 4 samples. When multiplexing retrospective samples, up to five libraries were sequenced simultaneously and sequencing for up to 24 hours was performed.

### Sequencing data processing

Base calling of raw data was performed using the manufacturer’ s proprietary software (guppy v3.1.5, CPU-based, fast mode, or v3.4.3, GPU-based, Oxford Nanopore Technologies, UK). Reads were then aligned to the hg19 human reference genome using minimap2 v.2.15 [9]. Copy number profiles were generated using R/Bioconductor and the QDNAseq package v.1.20 [14] using data from NA12878 reference DNA [5] for pseudo-germline subtraction.

Methylation status of CpG sites (5mC) was called using nanopolish v0.11.1 [16]. All workflows were implemented using snakemake v5.4.0 [8] for parallelization and deployment to a HPC cluster. When not using HPC, the pipeline was deployed to a single x86_64 workstation with 120 cores and 2TB RAM, augmented with an RTX2070 consumer-grade GPU (NVIDIA, Santa Clara, CA, USA) for base calling, running a Linux operating system (Ubuntu 18.04).

### Random forest classification

5mC signals at sites overlapping with sites probed by the Illumina BeadChip 450K array were then used to train a random forest classifier using the Heidelberg reference cohort of brain tumor methylation profiles [1]. Beta-values from the training set and sample set were binarized using 0.6 as a threshold value. When more than 50,000 features, the 50,000 most variable features were selected by standard deviation. Random forest classification was implemented in R using the ranger package v.0.12.1 [17]. The following parameters were modified from default: ntree = 20,000. Stratified sampling (to match the smallest class size, i.e., eight) was used to account for class imbalance. While random forests (RF) have been shown to be effective for many classification tasks, they typically report poor estimates of class probabilities. Given the importance of interpretable probability scores in a clinical context, the estimated class probabilities from such a classifier can be calibrated, which rescales predicted probabilities to be more accurately interpreted as confidence levels. We adopted a calibration strategy using Platt scaling by fitting a logistic regression model as previously described [11], as follows. In order to recalibrate the output of the RF classifier, probability reporting in the ranger function call was set to true. Platt scaling was originally suggested for binary classification tasks, so we reduced our multi-class classification task to a series of binary calibration tasks using the 1-vs-all method. Platt scaling was implemented by fitting logistic regression for each class, using the glmnet v.4.1 package in R. The recalibration was modelled on a binomial distribution.

### Prospective cases and cross-laboratory testing

As a pilot phase for a planned multicentric clinical trial (Universal Trial Number: U1111-1239-3456), a small series of prospective cases (N = 15) were evaluated regarding turnaround time and concordance to results from routine diagnostic workup. Prospective samples were obtained from the local neurosurgery department after written informed consent was given by the patient. Pseudonymized study data (start of surgery, time point of tissue receipt, DNA extraction, library preparation, sequencing metrics, classification results) was collected and managed using REDCap software [4], which was provided by the Berlin Institute of Health’ s Clinical Research Unit in a certified computing environment.

## Statistics

Data analysis was performed using R v.4.0.2. Receiver operator characteristics (ROC) were analyzed with the ROCit v.2.1.1 package. Figures were mainly visualized using ggplot2 v.3.3.2 and ComplexHeatmap v.2.6.2 packages [3].

## Code and data availability

The current nanoDx classification and analysis pipeline will be publicly available at https://gitlab.com/pesk/nanoDx (version v.0.2.1 was used for preprocessing of all sequencing data) upon publication. Source code for the outlined RF implementation and to reproduce all analyses and figures in this manuscript will also be available at https://gitlab.com/pesk/nanoBenchmark. Raw sequencing data from the discovery cohort will be deposited at the European Genome-phenome archive (accession tba). Methylation microarray raw data and methylation calls from the validation cohort will be deposited at ArrayExpress (accession tba).

## Results

### Robustness of pan-brain cancer classification using nanopore sequencing

In low-pass nanopore whole-genome sequencing, genome coverage is sparse and only a random subset of the ∼30 million CpG sites in the genome are probed. We have therefore proposed ad-hoc training of random forests (RF) using the overlap of the random CpG feature set of each sequencing run and the fixed feature space of the microarray-based training set [2]. Here, we used the Heidelberg brain tumor classifier as reference, which distinguishes 82 tumor entities and 9 non-tumor control classes. It implements a two-tier approach for methylation classes that are prone to misclassification without current clinical consequence (e.g., subtypes of IDH-wildtype glioblastoma). In these situations, superclasses, termed methylation class families (MCF), were defined followed by subtype classification where applicable. Similarly, we performed classification against a MCF level training set (composed of 8 MCFs and 67 methylation classes, totaling 75 classes) or the full training set of 91 methylation classes. Classification results were compared to the institutional WHO 2016 integrated diagnosis. The raw majority vote across the discovery cohort of 56 brain tumor samples was correct in 52/56 (92.9%) and 53/56 (94.6 %) cases for the MCF and full training set, respectively.

As a measure of the certainty of the classification, we calculated a pseudo-probability score by Platt scaling of RF raw vote counts. This transformation allows to apply a single cut-off value (or, synonymously, minimum score) to identify valid predictions. We used ROC analysis to identify the optimal cut-off value in the discovery cohort. For MCF level classification (75 classes), the calibrated score allowed reliable prediction of correct classification (AUC = 0.95, Fig. 1a) by applying a cut-off value of > 0.7 (which is slightly more conservative than the optimal cut-off of > 0.6 suggested by Youden index interpretation). This resulted in 88% sensitivity and 100% specificity in the discovery cohort (Fig. 1c). Youden index analysis suggested a highly similar cut-off (> 0.68) for the full methylation class-level (91 classes) training set (AUC = 0.79, Fig. 1b). Therefore, we decided to implement > 0.7 for both classification methods. This resulted in 62% sensitivity and 100% specificity to correctly predict methylation class in the discovery cohort (Suppl. Fig. 2a).

**Fig. 1.**
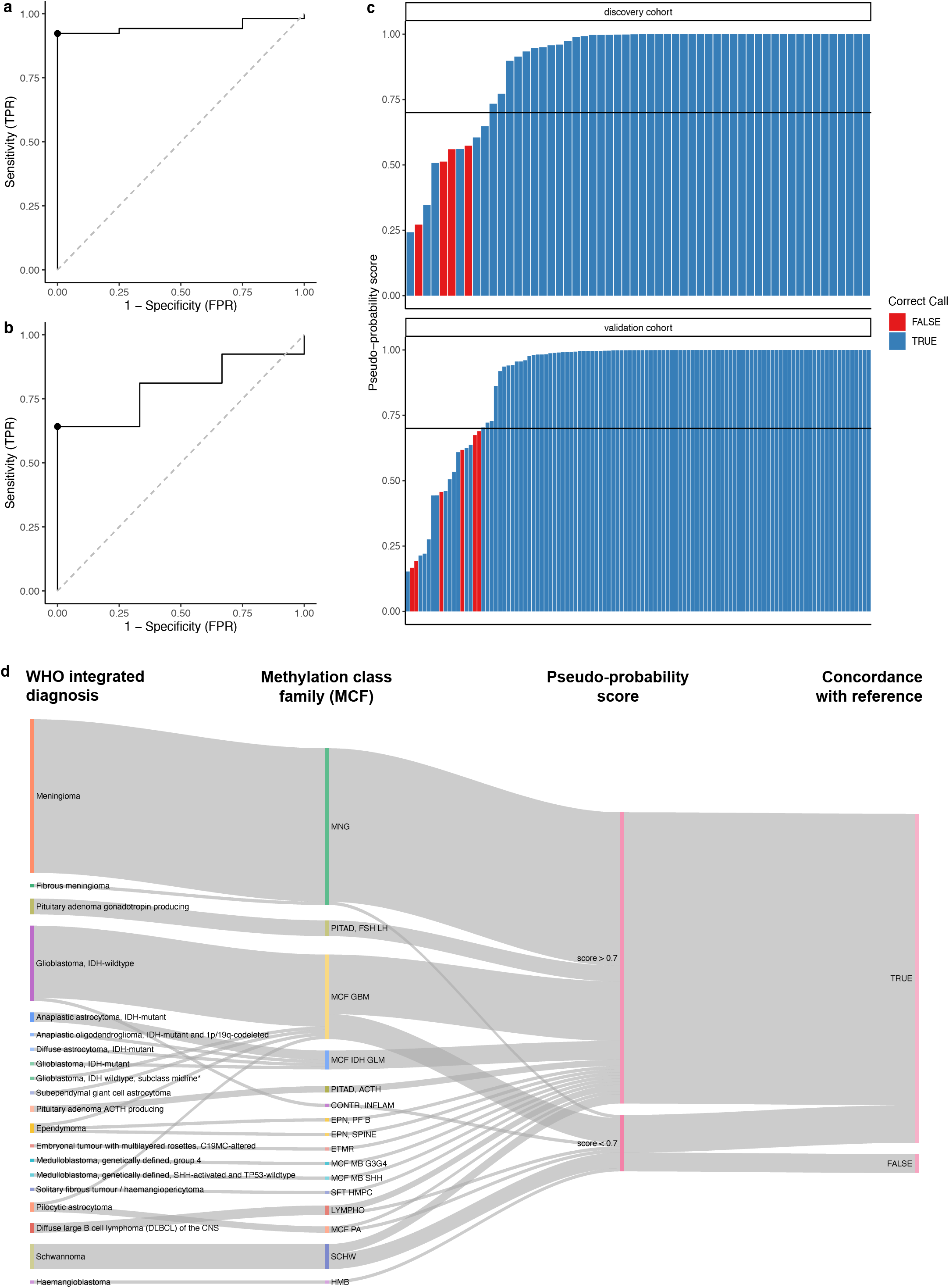
Performance of ad-hoc random forest pan-brain tumor classification. (a, b) Determination of cut-offs for pseudo-probability scores to detect correct classification using a methylation class family (a) and methylation class (b) training set. (c) Waterfall plots indicate relation of the methylation class family level classification result and the pseudo-probability rescaled classification score in the discovery (N = 56) and validation cohort (N = 111). Color indicates concordance (*blue*) or discordance (*red*) of the called methylation class family or methylation class with the institutional WHO 2016 integrated diagnosis. The cut-off identified by ROC analysis (> 0.7) is indicated by the horizontal solid line. (d) Classification results with respect to WHO diagnosis and corresponding methylation classification result in the validation cohort of N=111 independent samples.

**Fig. 2.**
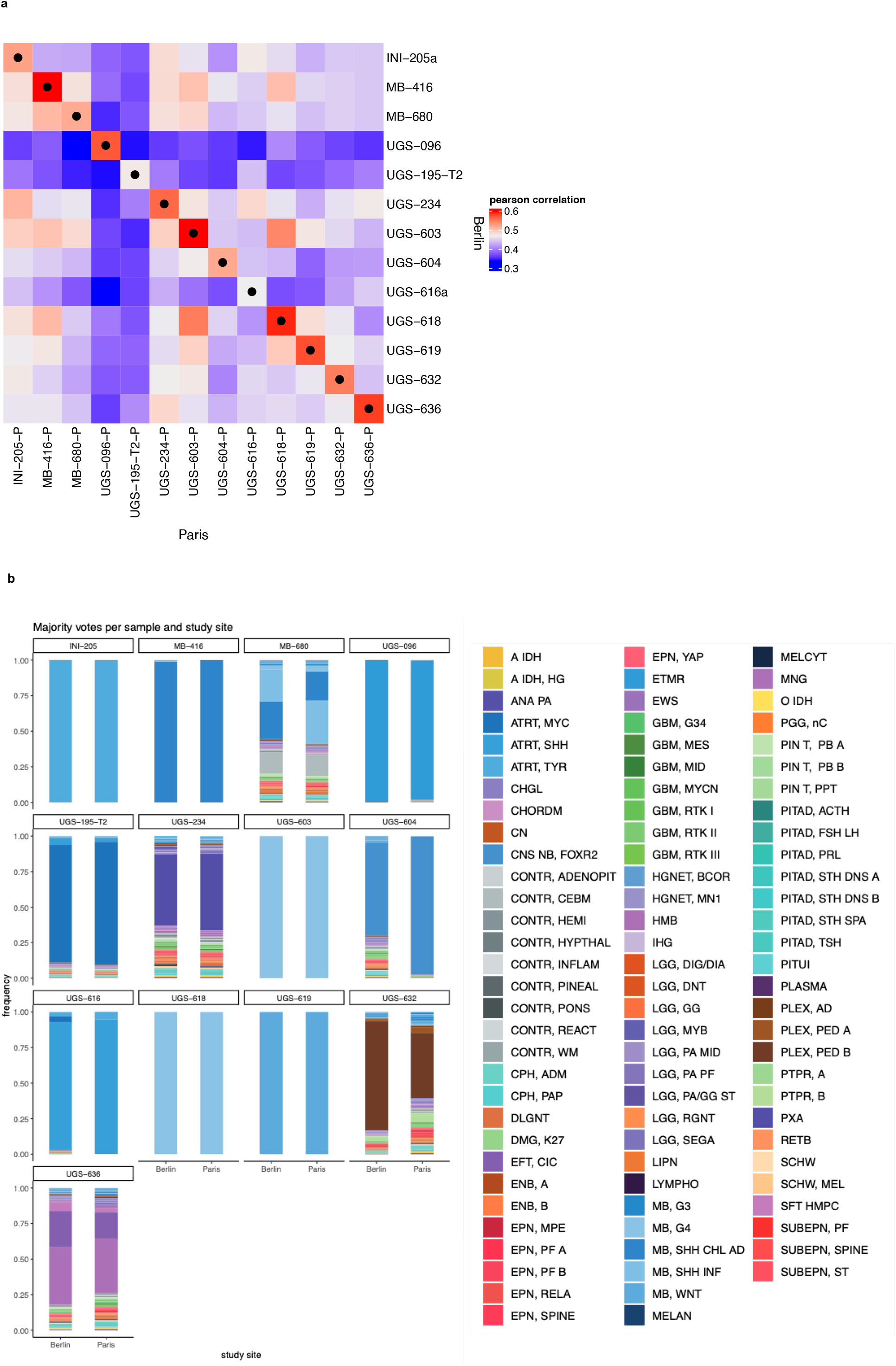
Cross-laboratory validation. (a) Pearson’s correlation of methylation status of shared CpG features between paired samples. Maximum correlation per row is indicated by black dots. (b) Comparison of random forest vote distribution for paired samples. Bar plots show recalibrated scores per methylation class.

We then applied this approach to an independent validation series of 111 primary brain tumor cases. On the MCF level, 105/111 cases (95 %) were classified correctly overall, i.e., a methylation class concordant with the sample’ s WHO integrated diagnosis was called. Applying a minimum score > 0.7 resulted in correct classification of all 93 cases with scores above the cut-off, corresponding to 89% sensitivity and 100% specificity (Fig. 1c, d). On the methylation class level, overall correct classification was found in 103/111 cases (93 %). Requiring a cut-off > 0.7 yielded 72% sensitivity and while retaining 100% specificity (Suppl. Fig. 2a, b). Matched array-based methylation data and classification results were available for 109/111 cases. Identical methylation classes were assigned in 89/109 cases overall and in 78/79 cases where a nanopore sequencing-based classification yielded a score > 0.7 (Suppl. Table 1).

### Impact of number of CpG features

Next, we investigated whether there was a correlation between the number of CpG features and recalibrated classification score as we hypothesized that a minimum number of features would be required for robust classification. First, we performed iterative random subsampling of CpG features for each sample in the discovery cohort. Second, we analyzed the out-of-bag (OOB) misclassification error of the training set and each sample’ s classification score with respect to the number of CpG sites used for RF training. Overall, no significant correlation between the number of CpG features and classification score was observed (Pearson’vs r = 0.02, p = 0.58). However, when less than 1,000 CpG sites were used, low scores were frequent and even misclassification with high confidence scores were observed (Suppl. Fig. 3). We therefore considered 1,000 overlapping CpG sites between the fixed feature space of the microarray-based training set and whole-genome nanopore sequencing the absolute minimum number for reasonable analysis.

### Cross-laboratory validation

Next, to assess reproducibility across laboratories, a series of N=13 tumors were profiled independently by nanopore whole-genome sequencing at two sites using the same DNA sample. Mean correlation of methylation calls of individual CpG sites over all 13 samples was 0.55 (SD = 0.046) and matched pairs always showed the best correlation across the series (Fig. 2a). Classification of matched samples was identical in 13/13 cases (Suppl. Table 2) with very similar raw vote distributions (Figure 2b). In addition, DNA methylation microarray (Illumina Infinium BeadArray 850K chip) data were available for 11/13 samples. Concordant results across all three tests were obtained in 10/11 cases. In the one mismatch case, the nanopore-based classification at both sites was concordant (Meningioma) but did not match the array-based classification (CNS Ewing sarcoma family tumor with CIC alteration).

### Benchmarking of nanopore sequencing over time

Nanopore sequencing data is generated sequentially over time and permits real-time analysis. Therefore, we next re-analyzed the discovery cohort by extracting subsets of sequencing data that were generated within a given time interval. As the raw yield is roughly proportional to elapsed sequencing time, the number of sampled CpG sites overlapping with the training set steadily increased over time. The minimum number of 1,000 CpG sites was sequenced within the first 30 min of the nanopore run in 39/56 samples (70 %, Fig. 3b). After 50 minutes, the correct call rate from cases within the discovery cohort obtaining a score > 0.7 and later correct classification result with their full set of CpG features was 100% (Fig. 3a). A correct, high confidence classification (i.e., correct call with score > 0.7) was made within 30 min of sequencing in 25/56 samples (45 %). Importantly, high confidence classifications based upon more than 1,000 CpG features, no matter at what time point they were made, were correct in 55/56 samples (Supp. Fig. 4).

**Fig. 3.**
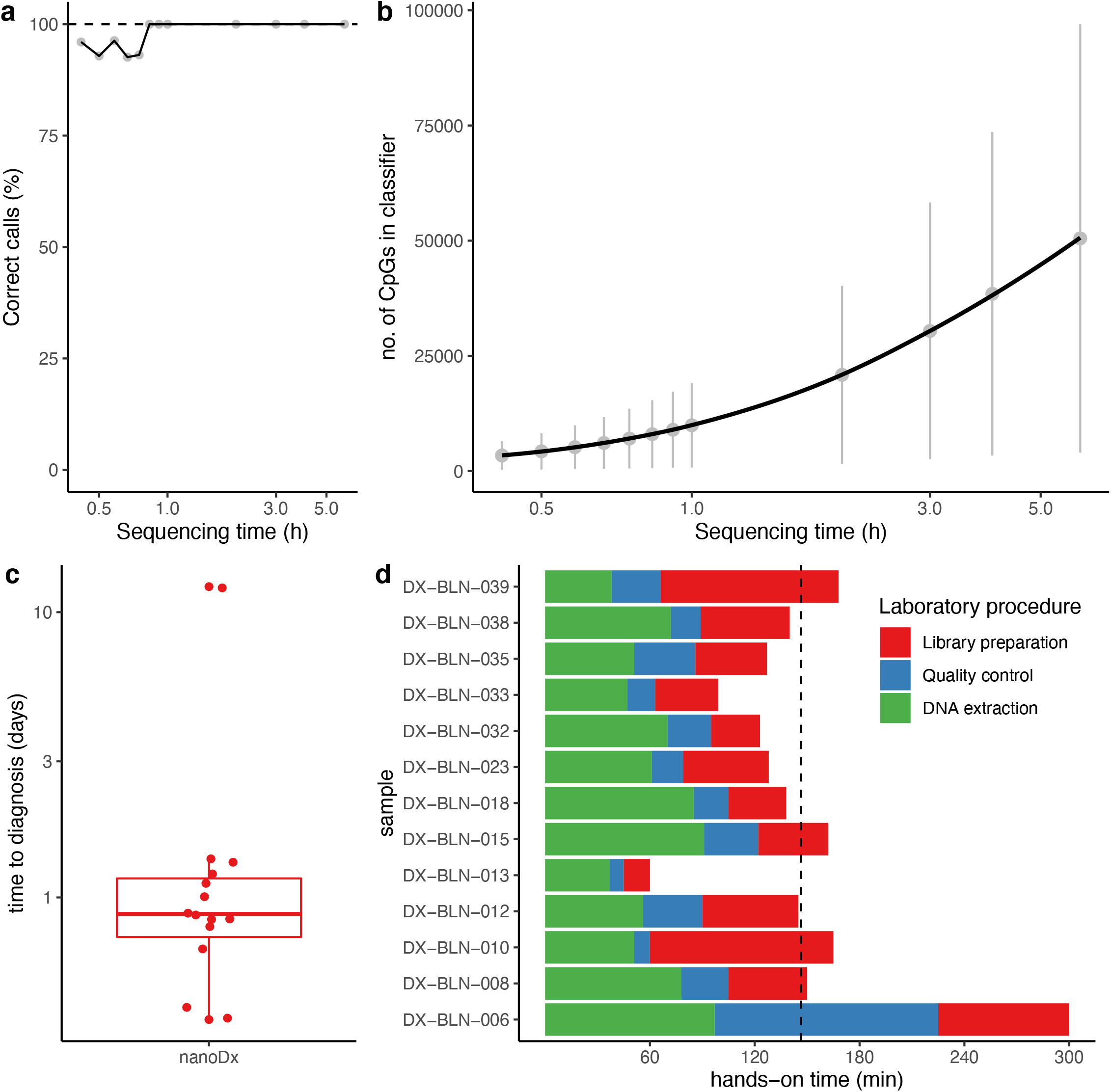
Benchmarking of nanopore sequencing over time and laboratory turnaround times. (a) Percent of correctly classified samples in the discovery cohort with a score > 0.7 with correct majority vote at given timepoint (in hours). (b) Number of CpG features with respect to sequencing time passed. Light grey lines show the standard deviation for each timepoint. (c) Boxplots indicate the median and quartile ranges of time elapsed from receipt of tumor material to timestamp of nanopore sequencing based classification reports, respectively. (d) Analysis of hands-on time with respect to time spent on DNA extraction (*green*), DNA quality control and quantification (*blue*) and library preparation (*red*). Average time elapsed across all samples is indicated as a dashed line.

### Benchmarking of turnaround times

Finally, as part of the pilot phase for a planned multicentric clinical trial (German Clinical Trials Register, Universal Trial ID: U1111-1239-3456), samples from N=15 patients were analyzed prospectively. Results of nanopore methylation-based classification were available in a mean of 39.4 hours (median = 21.1 hours) after receiving tissue from histological tumor purity assessment (Fig. 3c). Furthermore, hands-on-time spent on DNA extraction, quality control and nanopore library preparation was 146.5 minutes on average for singleplex sequencing (Fig. 3d).

## Discussion

In the present study, we demonstrate that methylation profiles generated by low-pass nanopore whole-genome sequencing allow robust and unbiased classification of primary brain tumors over the entire spectrum covered by the Heidelberg brain tumor classifier. We defined cut-off values for reliable clinical interpretation and show that ad hoc training and classification using random forests yields classification with very high specificity and acceptable sensitivity. Moreover, concordant and reproducible results across laboratories were obtained.

### Opportunities for nanopore methylation-based tumor classification

With a median time to diagnosis of 21 hours within the prospective cohort, nanopore-based methylation classification offers the chance to dramatically shorten turn-around times allowing completion of methylation-based molecular profiling without delaying first-line therapy. Nanopore methylation-based classification was usually available prior to immunohistochemistry in routine pathological workup due to delay by fixation and paraffin embedding. This allows neuropathologists to decide which stainings and complementary methods could further contribute to the diagnostic procedure, avoiding time-consuming sequential testing and accelerate the overall time to integrated diagnosis. Moreover, our data suggest further potential to reduce turnaround of nanopore-based testing and even intraoperative classification may be possible. In addition, genome-wide copy number profiles generated from nanopore WGS data have proven highly useful in diagnostic decision-making, replacing, e.g., time-consuming and costly fluorescence in situ hybridization assays.

### Machine learning aspects

Our current implementation of ad hoc random forests uses binarization of methylated allele frequencies to normalize for platform differences between microarrays (reference data) and nanopore sequencing. While this reflects all-or-nothing distribution found in promoters, epigenetic state encoded in regions with intermediate methylation, such as partially methylated domains, is likely lost. In addition, CpG sites called from individual long nanopore reads are not independent features but are currently treated this way. Exploiting this epi-haplotype information could possibly further improve classification.

### Limitations

While this study aims to offer a benchmarking of nanopore sequencing for DNA methylation-based classification of brain tumors and demonstrates its robustness, further research is needed in order to evaluate non-inferiority against current gold standard techniques (e.g., methylation bead array techniques) in larger cohorts. This will represent the scope of a planned multicentric clinical trial (German Clinical Trials Register, Universal Trial ID: U1111-1239-3456). In addition, while methylation bead arrays work well with formalin-fixated paraffin-embedded (FFPE) tissue, current protocols for nanopore sequencing depend on the availability of native or fresh-frozen tumor tissue. In order to support widespread implementation and analysis of archival tissues, protocols for robust nanopore sequencing from FFPE-derived DNA remain to be evaluated. However, with per-assay costs as low as $250 and initial hardware investment costs of $1000, a nanopore sequencing-based approach might be attractive in laboratories that see only a few neuro-oncological cases per week. Software compatibility with GPU-equipped multi-core PCs (in the range of $2000) significantly reduces cost for the compute infrastructure and eliminates the need to access a high-performance computation center.

In conclusion, we see great potential for a routine implementation of nanopore sequencing in DNA methylation-based classification in brain tumor diagnostics not only to shorten time to diagnosis but to augment neuropathological decision making and improve diagnostic precision. Further prospective evaluation in the context of a multicentric trial has just begun.

## Data Availability

The current nanoDx classification and analysis pipeline https://gitlab.com/pesk/nanoDx (version v.0.2.1 was used for preprocessing of all sequencing data) and the source code for the outlined RF implementation and to reproduce all analyses and figures in this manuscript https://gitlab.com/pesk/nanoBenchmark will be publicly available upon publication. Raw sequencing data from the discovery cohort will be deposited at the European Genome-phenome archive (accession tba). Methylation microarray raw data and methylation calls from the validation cohort will be deposited at ArrayExpress (accession tba).

## Table and figure legends

**Suppl. Fig. 1**

Study design and patient cohort. Tuning of machine learning parameters and ROC analysis to establish calibration score cut-offs were performed in a discovery cohort of N=56 samples. 13/56 samples were analyzed in two independent laboratories for cross-lab validation. 15/56 samples were collected and sequenced in a prospective fashion to benchmark turnaround times. Performance of the final classification approach was evaluated in an independent validation cohort of N=111 samples.

**Suppl. Fig. 2**

**Performance of ad-hoc random forest pan-brain tumor classification using the full Heidelberg reference set with 92 methylation classes**.

(a) Waterfall plots indicate relation of the methylation class (MC) level classification result and the pseudo-probability rescaled classification score in the discovery (N = 56) and validation cohort (N = 111). Color indicates concordance (*blue*) or discordance (*red*) of the called MC with the institutional WHO 2016 integrated diagnosis. The cut-off identified by ROC analysis (> 0.7) is indicated by the horizontal solid line. (b) Classification results (MCF, MC) with respect to tumor entity in the validation cohort of N=111 independent samples.

**Suppl. Fig. 3**

**Performance of classification with respect to number of CpG features**

Each sample in the discovery cohort (N= 56) was subjected to subsampling of a given number of random CpG features (ranging from 10 to 50,000 or the maximum number of features, if smaller) and underwent to ad-hoc random forest training and classification. Pseudo-probability scores are indicated with respect to the number of CpG features used for classification. Color indicates whether the RF methylation class family majority vote matched the correct diagnosis (blue) or not (red). The empirically identified cut-off value to accept the classification result (> 0.7) is indicated by a dashed horizontal line. 1,000 CpG features cut-off is represented by the vertical dashed line. Samples which had not been classified correctly with the full set of CpG features were excluded.

**Suppl. Fig. 4**

**Classification results over time**.

Each sample in the discovery cohort (N= 56) was re-classified after extraction of CpG features that were acquired within the indicated time interval. Color indicates whether the RF majority vote on methylation class family level matched the correct diagnosis (blue) or not (red). The empirically identified cut-off value to accept the classification result (> 0.7) is indicated by a dashed horizontal line. Samples with less than 1,000 CpG sites over the full sequencing time were excluded.

**Suppl. Table 1**

**In-depth information for each case in the discovery and validation cohort**

Supplementary table with additional information for each case in the validation and discovery cohort. The table contains clinical data, methylation-based classification results for the methylation class and methylation class family level as well as nanopore sequencing statistics for the discovery cohort.

**Suppl. Table 2**

**Cross-laboratory validation series**

Clinical data and methylation class results for all cases of the cross-laboratory validation cohort including methylation class calls for nanopore sequencing and Illumina EPIC 850K methylation array, if available.

## Acknowledgements

We thank Aydah Sabah for expert technical assistance. Computation has been performed on the HPC for Research cluster of the Berlin Institute of Health. N.B. and P.E. are participants in the BIH-Charité Clinical Scientist Program funded by the Charité – Universitätsmedizin Berlin and BIH. The project is funded by the The Brain Tumour Charity, UK.

